# A case-cohort longitudinal study for the analysis of microbial associations and viruses on the risk of celiac disease (MAVRiC)

**DOI:** 10.1101/2025.05.26.25328184

**Authors:** Kristian F. Lynch, Eric W. Triplett, Heikki Hyöty, Angelica P. Ahrens, Jutta E. Laiho, Joseph F. Petrosino, Richard E. Lloyd, Daniel Agardh

**Author notes:** **Corresponding author**: Kristian Lynch.

## Abstract

Celiac disease etiopathogenesis requires genetic predisposition and exposure to gluten before a child can develop the chronic autoimmune disorder. However, these factors alone are not sufficient. Even a child with persistent tissue transglutaminase autoantibodies (tTGA), i.e., celiac disease autoimmunity (CDA), does not necessarily develop celiac disease. Larger longitudinal studies are needed to determine the impact of time-varying infections and gut microorganisms on the subsequent and specific risk of celiac disease. The aim was to design a celiac disease case-cohort longitudinal study using The Environmental Determinants of Diabetes in the Young (TEDDY) study.

Following the TEDDY cohort up age 3-years, CDA was confirmed in 704 of the 6132 genetically at-risk children. Celiac disease onset was defined as the age CDA developed when followed by a biopsy-proven diagnosis. A competing risk analysis was performed on celiac disease onset cases (CD-onset) as well as CDA children with no diagnosis (CDA-only) and results show genetic factors (additional HLA-DR3-DQ2 haplotype, higher non-HLA polygenic risk score, sex-girl) and a more rapid increase in gluten-consumption correlate with significant increased risk of both outcomes. However, reports of virus-related respiratory infections in August to October during follow-up correlated with an increased risk of a CD-onset but not with CDA-only. To create a case-cohort study, a sub-cohort of 561 children (9% sampling fraction) was first randomly selected to represent the TEDDY cohort over time while at risk of CDA. It included 483 children followed until age 3-years and 78 children followed before developing CDA (CDA-only n=41/78, CD-onset n=37/78). All incident CD-onset children (N=306) were included to form the case group. This case-cohort will be utilized to analyze virus antibodies and bacteriome from longitudinal plasma and stool samples (the Microbial Associations and Viruses on the Risk of Celiac disease study, MAVRiC).

## Introduction

Celiac disease is a complex, primarily T-cell, immune mediated^1^ systematic disorder that manifests due to damage caused to the lining of the intestinal mucosa in the small bowel^2^. Without screening, absorption of nutrients is often impaired, leading to malnutrition and possibly both gastrointestinal and non-gastrointestinal symptoms. Globally, the prevalence of celiac disease exceeds 1.4% with an increasing trajectory and an impact that is highest for the child^3^.

The age of development of celiac disease in childhood is highly variable as revealed by cohort studies. The onset of disease is known to require two components, the ingestion of gluten to elicit an autoimmune response^4^ and the presence of HLA-DR3-DQ2 or DR4-DQ8 haplo-genotypes. While additional genetic contributions come from family history, female sex, and non-HLA loci, only 50% of the risk can be explained by genetic variation and gluten ingestion is not sufficient for celiac disease to occur. Epidemiological studies strongly implicate a role of environmental factors such as infections and gut microbiota in the triggering or acceleration of disease development. However, the identification and characterization of microbial associations is difficult without studies that frequently collect biological samples and without a defined longitudinal outcome indicating when mucosal damage is truly initiated over time. A biopsy has been considered the “gold standard” for diagnosis, but a positive histology alone is not sufficient and a final diagnosis should include a combination of positive serology and histology, with or without classical clinical features and a response to a gluten free diet (GFD) ^5^. These time varying criteria are unlikely to be measured continuously over time or even develop simultaneously. Nevertheless, cohort studies consistently show that peak incidences of both CDA seroconversion and celiac disease diagnosis occur after 1-year and before school age. Thus, the prodromal period between positive serology and histology is short enough at this young age to link disease initiation close to CDA seroconversion. The early peak incidences further suggest that environmental factors may act early in life to promote inflammation, reduce tolerance, or condition an aggressive immune response to the rising gluten consumption. The study of incident CDA by age 3 years with a subsequent biopsy-proven diagnosis offers the best opportunity to study factors such as early viruses and bacteria as candidate triggers of the onset of celiac disease.

The Environmental Determinants of Diabetes in the Young (TEDDY) prospective study^8,9^ has screened CDA annually and followed up for celiac disease among HLA-DR3-DQ2 or HLA-DR4-DQ8 risk children up to 15 years of age at six clinical sites in four countries across two continents. TEDDY has identified over 113,000 reported infectious episodes in early childhood while at risk of CDA and has reported correlation between CDA with reported gastro-intestinal infections (GIE), season of birth and enteroviruses. Additionally, TEDDY has collected monthly stool samples and quarterly plasma samples from all children, between 3 and 48 months of age, making it possible to study whether viruses and/or bacteria are associated with subsequent disease risk.

The primary aim of this study was to select a longitudinal outcome defining celiac disease onset using the TEDDY study and design an ancillary study, **M**icrobial **A**ssociation and **V**iruses on the **Ri**sk of **C**eliac disease (MAVRiC), to examine the role of bacterial and viruses with risk of celiac disease. A secondary aim is to re-examine reported infections in the TEDDY cohort with the risk of CD-onset to generate hypothesis as to the time-dependent role of microorganisms on risk of celiac disease.

## Methods

### Study Designs

#### Cohort study (TEDDY)

As an observational birth cohort study, TEDDY is following children for type 1 diabetes (T1D) until 15 years of age. The study aims to identify environmental risk factors of islet autoimmunity (IA) and CDA as primary outcomes and will follow for T1D and celiac disease as secondary outcomes. Between September 2004 and February 2010, 424,788 newborns across three clinical centers in the US (Colorado, Georgia/Florida, and Washington state) and three centers in the EU (Sweden, Finland, Germany) were screened for HLA-DR-DQ haplo-genotypes and 21,583 children were deemed eligible. At the age of 3 to 4 months, 8,676 of these families enrolled in the study of which 6,555 (76%) participated in annual screening for CDA by age 3-years. Nearly all enrolled children (96%) had one of the following four HLA-DR-DQ haplo-genotypes: DR3-DQ2/DR3-DQ2 (20.7%), DR3-DQ2/DR4-DQ8 (39.0%), DR4-DQ8/DR4-DQ8 (19.5%), DR4-DQ8/DR8-DQ4 (16.7%). Additional characteristics at the time of enrollment are shown elsewhere. The follow-up of children consisted of quarterly clinical visits and blood-draws until the age of 48 months. Stool samples were collected monthly starting at enrollment until age 48 months. To persevere priority samples, children positive for IA before 4 years of age (n=423/6555) were excluded at the request of the TEDDY study. The final Celiac Cohort study included 6132 children. This ancillary study was approved by the TEDDY ancillary committee and is being led by several members of the TEDDY Infectious Agents and Celiac Disease committees.

#### Case-Cohort study (MAVRiC)

To combine the flexible advantages of a cohort study with the efficiency of a nested case-control (NCC), a case-cohort design was chosen for the MAVRiC study to address the role of virus infections and bacteriome in the development of celiac disease in the TEDDY study. Unlike an NCC study that involves first identifying incident cases during follow-up and then selecting 1 to 3 controls to match with each case, the case-cohort design involves first selecting a sub-cohort of children from the original cohort as a representative baseline population. All incident cases outside this sub-cohort are later included to complete the case-cohort study design. Cases chosen into the sub-cohort by random chance represent time-varying controls while they remain at risk of the case event. They are weighed differently in analysis to account for the fact that cases are oversampled. The selection of the sub-cohort allows for its flexible re-use as a comparison group both in stratified analysis and if the study needs to be extended in the future to include additional cases.

The sub-cohort consisted of 600 randomly selected children from the original cohort of 6555 (9.2% sampling fraction) before IA positive cases were excluded. Of the children selected, 39/600 developed IA before 4-years of age and were omitted from the remaining analysis. The random selection was performed before excluding the IA cases so that sensitivity analyses could be performed at a later stage to study the impact of their exclusion.

### Onset of celiac disease as a primary outcome

The inclusion into the MAVRiC case-cohort study of all children developing any CDA was cost prohibitive. Moreover, any biopsy conducted to determine celiac disease was often a year after CDA development and after the disease had likely developed. The primary outcome of interest in this study was the onset of celiac disease (CD-onset) defined as the age when CDA develops before age 4-years and celiac disease is subsequently confirmed with a positive biopsy. Children developing CDA without a subsequent celiac disease diagnosis (CDA-only, n=398) were examined in the full cohort only and were not included in the MAVRiC case-cohort study. A comparison of the outcome definitions in TEDDY and MAVRiC studies are presented in **Table 1**.

**Table 1.** Outcomes celiac disease autoimmunity (CDA) and celiac disease in the TEDDY cohort and case outcomes of interest in the MAVRiC case-cohort.

| Outcomes in the TEDDY Celiac cohort | Definition and time of onset | N <sup>a</sup> |
| --- | --- | --- |
| Any CDA<br>(Primary outcome) | Two consecutive positive tTGA results (level >1.3 U/ml) at least 3 months apart. Time of onset was age of seroconversion (first positive sample). | 704/6132 |
| Celiac disease<br>(Secondary outcome) | An intestinal biopsy showing a Marsh score 2 or greater during follow-up (n= 283). Time of onset at age of positive biopsy.<br>No biopsy performed. Child had CDA and max average tTGA levels for any two consecutive samples during follow-up was ≥100 U/ml indicating serological proven celiac disease with >99% specificity for biopsy proven celiac disease (n= 23). Time of onset was at age of first sample. | 306/704 |
| **Case outcomes of interest for the MAVRiC study** |  |  |
| CD-onset <sup>b</sup><br>(primary outcome) | Any CDA before age 4-years followed by a celiac disease diagnosis. Time of onset at age of CDA seroconversion (first positive sample) | 306/704 |
| CDA-hi <sup>b</sup><br>(specific outcome) | Any CDA before age 4-years with tTGA level of the persistent sample ≥60 U/ml indicating serological proven celiac disease with >90% specificity for biopsy proven celiac disease within a year of CDA development. Children with (n=182/306) and without (n=20) a subsequent celiac disease diagnosis are included. Time of onset was age of CDA seroconversion (first positive sample). | 202/704 |
| **CDA children not selected as a case in the MAVRiC study** |  |  |
| CDA-only <sup>b</sup> | Any CDA before age 4-years with no subsequent celiac diagnosis during follow-up in TEDDY. Onset at age of CDA seroconversion | 398/704 |

<sup>a</sup>= N refers to the number of children developing CDA before age 4-years of age (n=704)
<sup>b</sup>= Any CDA cases not included in the outcome definition were treated as a competing risk in analysis and are censored at the age of CDA seroconversion (i.e. they still serve as controls and are representative of the cohort in prior risk sets).

The TEDDY protocol was designed for timely and thorough ascertainment of the development of CDA and subsequent celiac disease. The annual screening for tTGA started at 24 months of age and children positive for tTGA had their subsequent sample tested to determine if the child had developed CDA. Two consecutive positive tTGA results at least 3 months apart defined any CDA. If a child missed their annual visit or had no sample available, tTGA screening results were collected at the next visit. Any child positive for tTGA had their remaining available samples tested retrospectively to determine CDA seroconversion. Nearly all the children developing any CDA up to the age of the 27-month visit tested positive on their first annual screen, while CDA between 30- and 48-month visits were initially screened negative. The incidence proportion of any CDA by age 3-years in the TEDDY Celiac Cohort was 11.5% (704/6132). Tissue TGA was monitored frequently in the entire cohort after CDA seroconversion, with prompt referral of those positive for further medical evaluation. An intestinal biopsy showing a Marsh score 2 or greater was considered to have biopsy-proven celiac disease.

In all, 306 of the 704 children developing any CDA before 4 years of age were confirmed with celiac disease before age 14-years or August 31^st^, 2020, whichever came first. This was the primary outcome of the MAVRiC study. Because the identification of CD-onset was determined through retrospective analysis of the TEDDY cohort, it was necessary to have a secondary CDA phenotype (CDA-hi) that indicates a higher specificity for mucosal damage during CDA development and not after (**Supplemental Figure 1**). This was to ensure microbial correlations found with risk of CD-onset also occurred with early evidence of disease progression at the time of CDA development and not on factors occurring only after CDA development. Tissue TGA patterns during CDA development were examined for correlation with a subsequent celiac disease diagnosis. All but seven of the 704 children were followed in TEDDY after CDA development. The median (IQR) number of years from the first sample positive for tTGA positive to the diagnosis of biopsy-proven celiac disease was 1.1 (0.8, 1.7) years. Compared to the first positive sample, the tTGA level of the persistent positive sample more clearly correlated with celiac disease diagnosis (**Supplemental Figure 2**). Furthermore, tTGA level distributions of the persistent positive sample compared to the first positive samples showed stronger correlation (r=0.86, p<0.001) with the child’s maximum average tTGA levels across any two consecutive samples during following (r=0.54, p<0.001, **Supplemental Figure 3**). Of the children with CDA-hi (n=202) with tTGA level ≥60 U/ml in the persistent positive sample, 10% (20/202) were not subsequently determined for celiac disease (i.e. CD-onset). These 20 CDA-only children were still added to the MAVRiC study as a baseline for secondary outcome analysis using only serological information; 11/20 had a follow-up biopsy that was negative and 9/20 had max average tTGA level in follow-up samples between 40 and 100 U/ml with no biopsy.

### Infectious episodes

Symptoms or diagnosis of any illnesses were recorded by the parents in a TEDDY diary-book or at each clinic visit. Infections reported from the previous visit were collected by study nurses who translated illness reports into diagnosis codes according to the ICD-10 classification. Infectious disease data processing and categorization in the TEDDY used an infectious episode (RIE) approach, which reduces the possibility of overestimation of reported exposure due to multiple symptoms and/or diagnosis reports during a single microbial infection. In the present study, four common categories of respiratory infectious episodes were of interest: 1) virus related respiratory type of infections that included mostly of common colds but also laryngitis and tracheitis, specific indication of influenza, enterovirus, chicken pox (varicella), measles, mumps, rubella herpes simplex virus infection and other virus infections not elsewhere classified; 2) infections of ear and mastoid process; 3) bronchitis and lower respiratory infections and 4) conjunctivitis. Two common categories of GIE were 1) gastroenteritis with report of infection and 2) gastrointestinal symptoms. All infections and categories were identified as having a record of an ICD-10 code since the last scheduled clinic visit. Given the season of birth and enteroviruses was of particular interest, the common virus-related respiratory infection was examined by a 3-month season starting with infections reported in the fall between August and October when enterovirus prevalence was expected to peak (**Figure 1A**).

**Figure 1.**
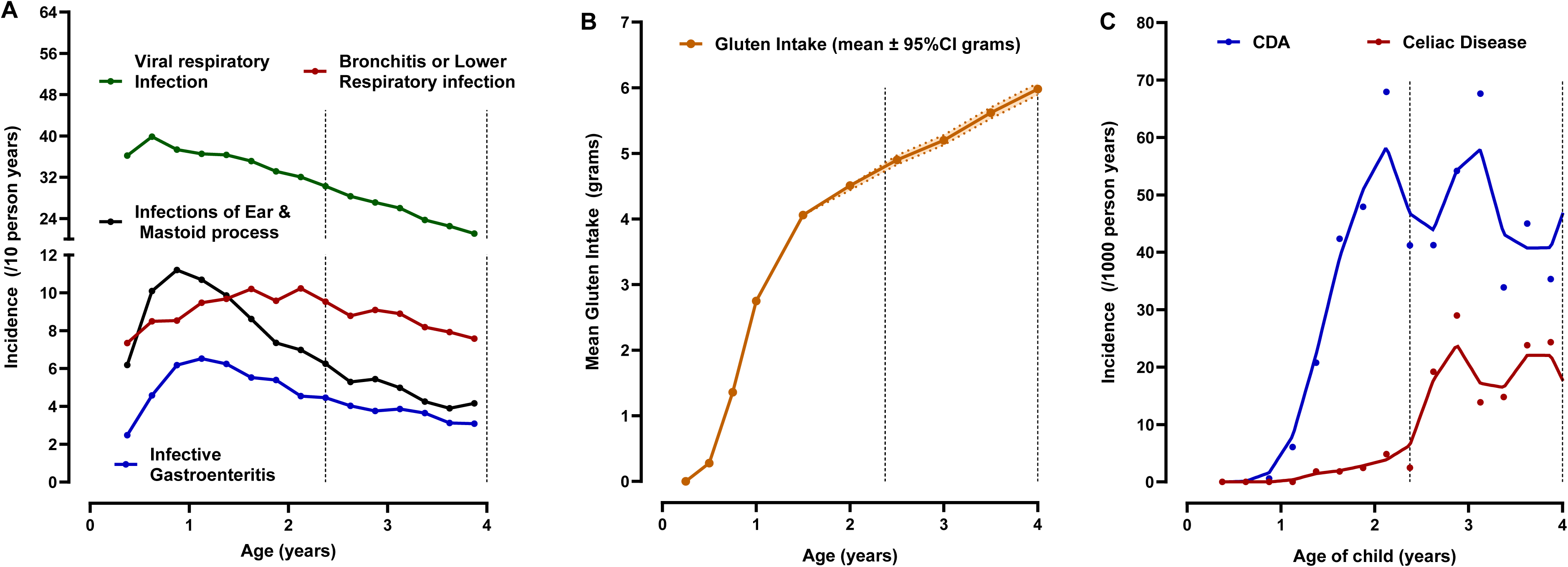

***Gluten intake*** was estimated from 3-day food records collected at ages 6, 9, and 12 months and biannually thereafter. TEDDY has previously reported on the gluten intake during the first 5 years of life. Higher intake correlates with an increased risk of CDA and celiac disease, whereas the age of introduction to gluten is not associated with these outcomes. From the age of 6 until 18 months, gluten intake rose rapidly (**Figure 1B**). Thereafter, the rate of increase was more gradual. Since only 44 CDA cases were observed before the age of 18 months (**Figure 1C**), the rapid rise in daily intake was captured by a linear slope trajectory between the age of 6 and 18 months while at risk of CDA and was calculated by fitting a linear mixed model with a random slope. If a child missed a 6-month visit, gluten intake at the 3 month-visit was included. The extracted slopes describing the rise in gluten intake for each child were accounted in every correlation model examining reported infections with risk of CDA. The average slope for a child during this age year was +3.8 per average daily grams. Child on a gluten free diet was rare before CDA development and common after CDA development (**Supplemental Figure 4)**.

***A polygenic risk score*** (PRS) for celiac disease was created to account for genetic risk outside the HLA region. Any non-HLA single nucleotide polymorphisms (SNPs) previously listed associated with CDA or celiac disease in TEDDY and available from the ImmunoChip or TEDDY-T1DexomeChip data were studied for their association with CD-onset before the age of 4 years. Each SNP was examined in relation to the risk of CD-onset using Cox proportional hazard models adjusting for country of residence, sex, HLA-DR-DQ haplo-genotype and the first three principal components describing ancestry. The PRS was created that summed the product of log beta coefficient (log hazard ratio=log HR) from the proportional hazard model and number of minor allele SNPs **(Supplemental table 1)**. The polygenic score was included in the final model to account for non-HLA loci.

### Statistical analysis

A competing risk analysis of CD-onset, CDA-hi and CDA-only was performed on the whole cohort (n=6132). Multivariate proportional hazard models were used to characterize associations with genetic risk factors, sex, demographic factors, exposure to gluten and infectious episodes all of which were previously reported associated with any CDA or celiac disease. All children developing CDA were censored at the time of seroconversion including the CDA group of interest and competing CDA groups. Effect sizes were described by outcome-specific hazard ratios with 95% confidence intervals. Categories of infectious episodes were examined as time-invariant factors during the first year of life and as time-varying factors during any year of follow-up. The age of infection was modeled as a step function and infections during follow-up prior to CDA were modeled as a lag function. The same analysis on CD-onset was repeated in the MAVRiC case-cohort study. However, CD-onset (and CDA-hi) cases were intentionally over-represented in a case-cohort design which necessitated an appropriate adjustment to the proportional hazard models. Instead of using the partial likelihood, a pseudolikelihood estimator was implemented. This approach incorporates weights to produce estimates of hazard ratios comparable to those observed in the cohort. Prentice weights were used to correct for oversampling of cases in the case-cohort design and confidence intervals were calculated using robust standard errors. Unless otherwise stated, p-values less than 0.05 were considered significant. All statistical analyses were performed using R, version 4.4.2 (www.R-project.org), SAS, Version 9.4 (SAS Institute Inc. Cary, NC, USA), and figures generated using GraphPad PRISM 5.03 (GraphPad Software Inc., Sand Diego, CA).

## Results

### Factors with specific risk for CD-onset in the whole cohort

Factors previously reported associated with any CDA in TEDDY up to age 3-years, and summarized in a review, were examined together in a multivariate analysis on the specific risk of three CDA groups; CD-onset, CDA-hi and CDA-only. Female-sex, number of HLA-DR3-DQ2 haplo-genotypes, first degree relative with celiac disease, an increase in the non-HLA PRS, a rapid rise in gluten intake between 6 and 18 months, born in the late spring or summer compared to other months were independently associated with an increased risk of CD-onset (**Table 2**). In addition, the number of virus-related respiratory infectious reported August to October during first year of life correlated with a significant increased risk of CD-onset (/additional fall RIE, HR=1.28, 95%CI = 1.12 – 1.48, p=0.0005). After accounting for these factors, the country of residence, increase in fiber intake during the first year, mother’s education, age mother stopped breastfeeding, age child started gluten were not associated with risk of CD-onset (not all data shown in **Table 2**). Except for family history of celiac disease, all the factors correlating with CD-onset also correlated significantly with increased risk of CDA-hi that was highly predictive of a celiac diagnosis shortly after CDA seroconversion.

**Table 2.**
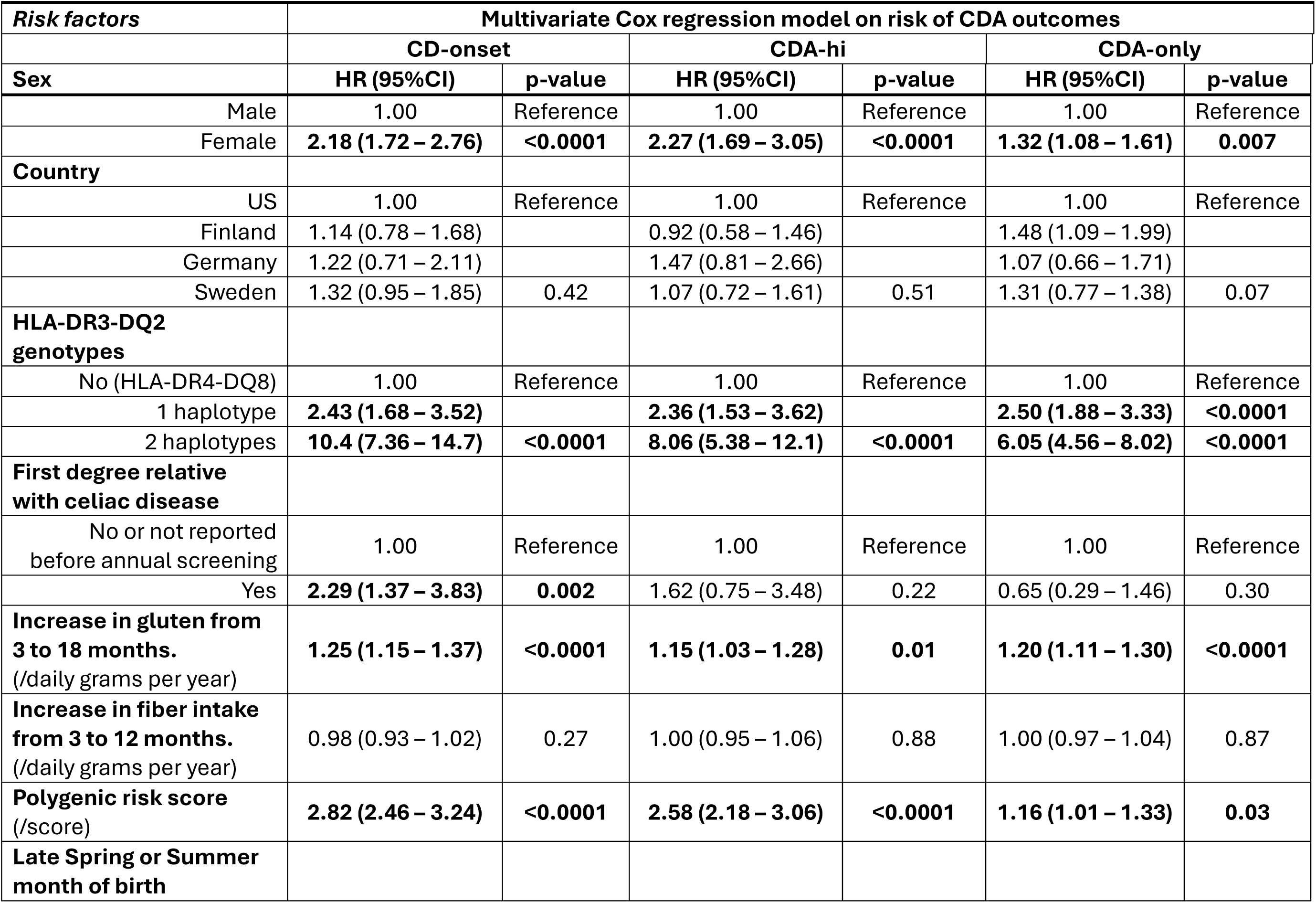
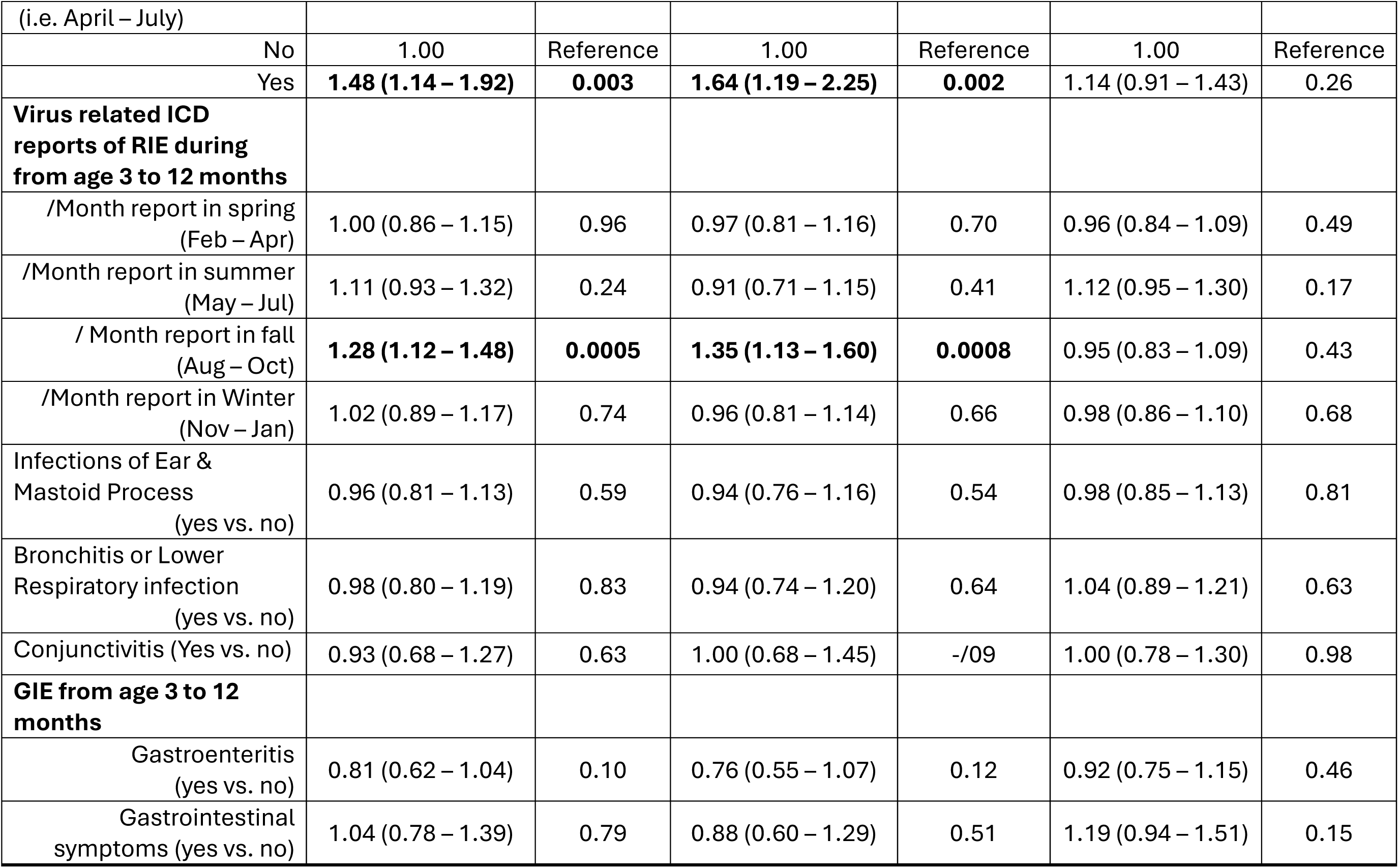
Cox regression modeling on risk of CD-onset, CDA-hi and CDA-only before age 4-years in the TEDDY Celiac Cohort.

However, it was noted that the risk of CD-onset and not CDA-hi, (i.e. CDA development with tTGA levels <60U/ml in the persistent sample and subsequent biopsy proven celiac disease) was more strongly correlated with a family history of celiac disease (HR=2.93, 95%CI = 1.46 – 5.88, p=0.002), a more rapid rise in gluten intake between 6 and 18 months (HR = 1.43, 95%CI = 1.26 – 1.63, p<0.001), and the risk was marginally reduced with a 3 to 12 month rise in fiber intake (HR=0.93, 95%CI = 0.87 – 1.00, p=0.07). Birth month and virus-related RIE reports in the fall during the first year of life did not correlate with this CDA group. Similarly, a late spring to summer month of birth and RIE reports in any season during the first year of life were not associated with the risk of CDA-only (i.e. risk of CDA with no subsequent celiac disease diagnosis, **Table 2**).

### Age and timing of infectious episodes on risk of CD-onset in the whole cohort

The impact of RIEs and GIEs overall and by category on the risk of CD-onset was examined in more detail. In any year during follow-up, RIEs but not GIEs correlated with a subsequent increased risk of CD-onset (additional infection, HR=1.14, 95%CI = 1.02 – 1.26 p=0.02). An examination of the common category of RIEs showed more reports of virus-related respiratory infections during the fall (August to October) in any year during follow-up explained this increased risk of CD-onset (/additional fall infection, HR=1.24, 95%CI =1.08 – 1.43, p=0.002). Given the similar result with fall infections during the first year, **Table 2**, analysis stratification analysis by age of CD-onset was conducted (<2.5 years vs. 2.5 - <4.0 years). The age ranges were indicative of whether the child was identified with CDA on their first annual screened for CDA at age 2-years or in later years.

Each additional virus related infections reported in the fall in the first year correlated with an increased risk of CD-onset before 2.5 years (additional fall infection, HR=1.56, 95%CI = 1.30 – 1.88, p<0.0001), but not from 2.5 to <4years (/additional fall infection, HR=1.02, 95%CI =0.81 – 1.27, p=0.89, age*interaction p=0.007). No other reported infectious episode categories were associated with CD-onset before or after 2.5 years.

These infection reports were examined in greater detail between quarterly visits on the risk of CD-onset before 2.5 years. Virus-related RIE reported during the fall in the first year of life correlated with strong consistency both by fall month and by quarter age (3 to 6, 6 to 9 and 9 to 12 months) on the risk of CD-onset, **Figure 2**.

**Figure 2.**
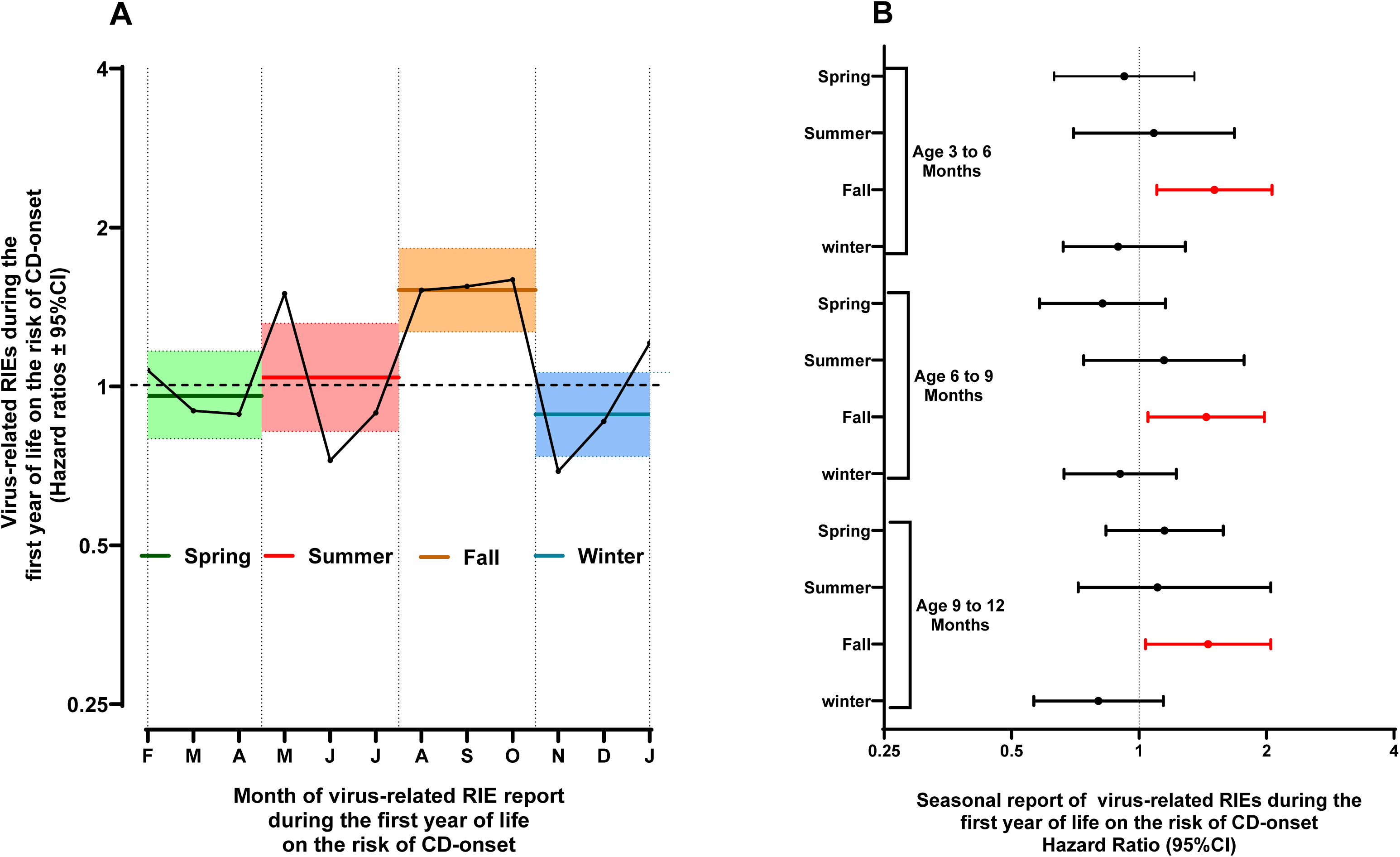

### The MAVRiC CD-onset case cohort study

The sub-cohort random selection from the TEDDY Celiac Cohort included 483 children who were followed in TEDDY through age 3-years, and 78 children who were followed until they developed CDA. By chance, these children were chosen to serve as time varying controls representative of the whole TEDDY cohort over time before their seroconversion. The inclusion of CDA children will allow for analysis of factors associated with the hazard-risk of disease. The representative CDA group included 37 children who were subsequently developed CD-onset by age 3-years and 41 children with CDA-only, **Figure 3**. All incident CD-onset cases are included in the MAVRiC case-cohort designed study. A description of the sub-cohort and TEDDY celiac cohort by country, sex, HLA, CD-onset and by CDA-only are shown in **Table 3**. Similarly, the 202 CDA-hi cases are characterized by country, sex and HLA, **Table 4**.

**Figure 3.**
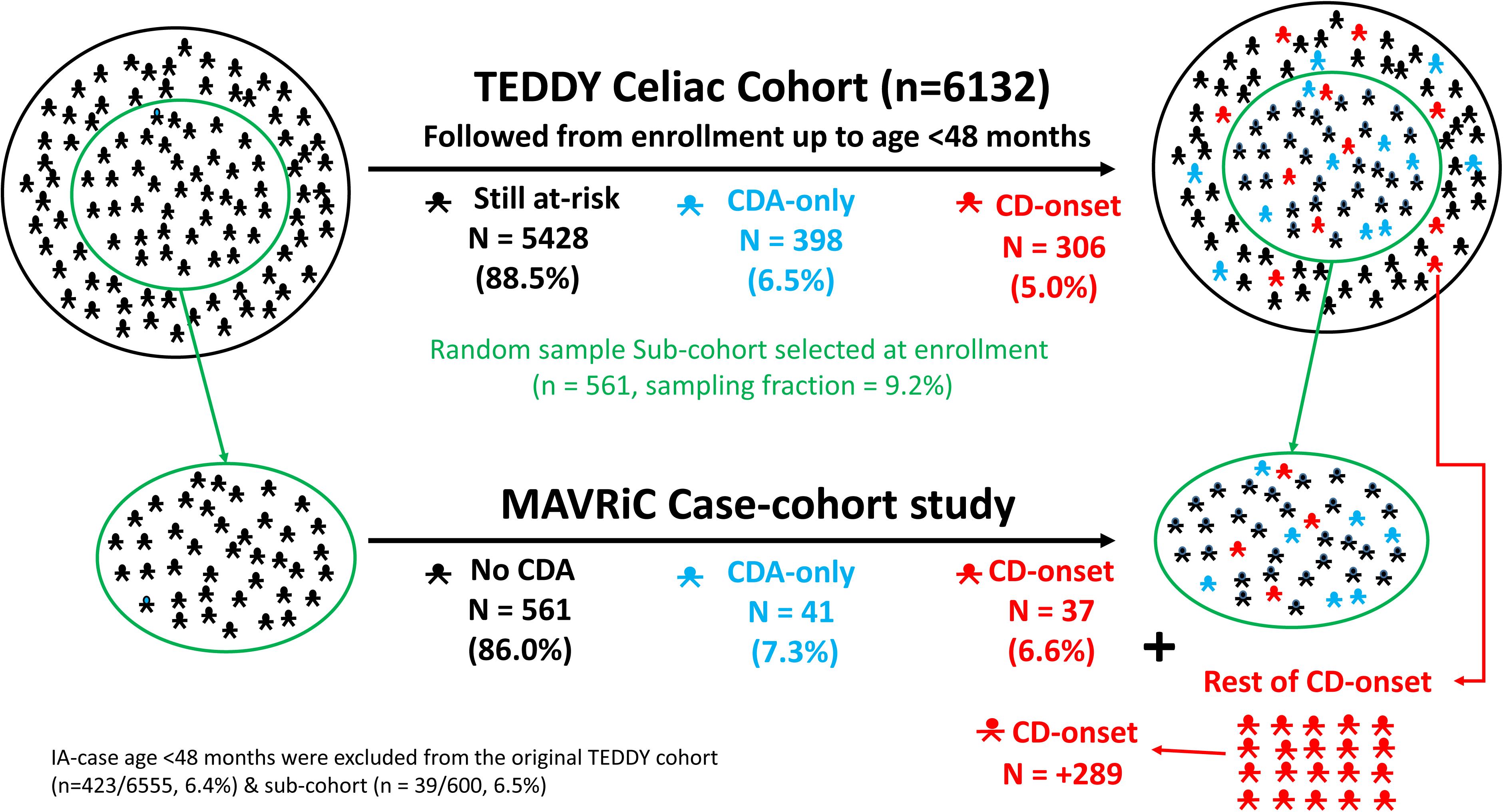

**Table 3.**
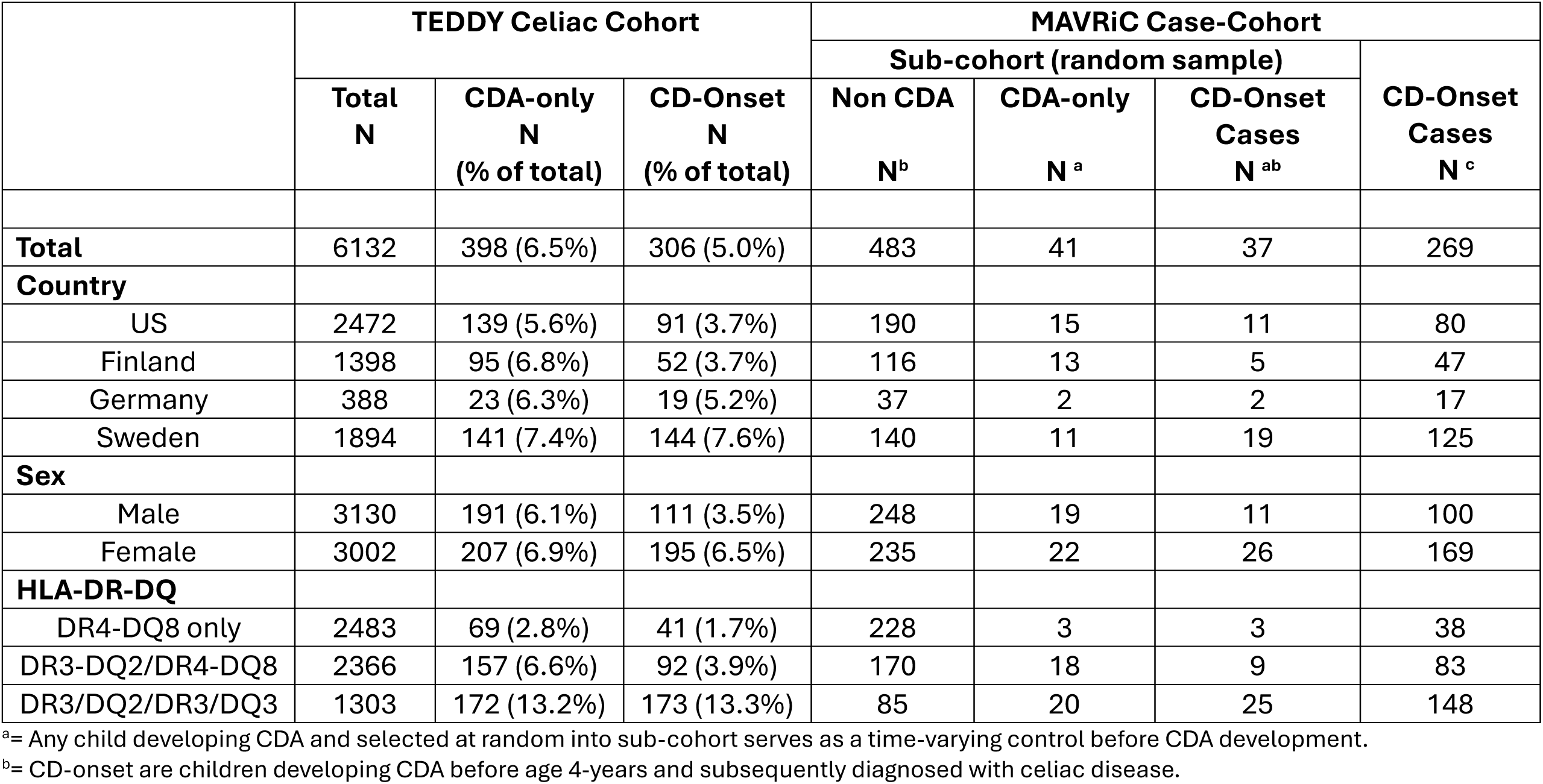
The MAVRiC Case Cohort study consisted of a sub-cohort (n=561) randomly chosen from the TEDDY Celiac Cohort and the inclusion of all children with CD-Onset ^b^ before 4 years of age.

|  | TEDDY Celiac Cohort |  |  | MAVRiC Case-Cohort |  |  |  |
| --- | --- | --- | --- | --- | --- | --- | --- |
|  | Total<br>N | CDA-only<br>N<br>(% of total) | CD-Onset<br>N<br>(% of total) | Sub-cohort (random sample) |  |  | CD-Onset<br>Cases<br>N <sup>c</sup> |
|  |  |  |  | Non CDA<br>N<sup>b</sup> | CDA-only<br>N <sup>a</sup> | CD-Onset<br>Cases<br>N <sup>ab</sup> |  |
| **Total** | 6132 | 398 (6.5%) | 306 (5.0%) | 483 | 41 | 37 | 269 |
| **Country** |  |  |  |  |  |  |  |
| US | 2472 | 139 (5.6%) | 91 (3.7%) | 190 | 15 | 11 | 80 |
| Finland | 1398 | 95 (6.8%) | 52 (3.7%) | 116 | 13 | 5 | 47 |
| Germany | 388 | 23 (6.3%) | 19 (5.2%) | 37 | 2 | 2 | 17 |
| Sweden | 1894 | 141 (7.4%) | 144 (7.6%) | 140 | 11 | 19 | 125 |
| **Sex** |  |  |  |  |  |  |  |
| Male | 3130 | 191 (6.1%) | 111 (3.5%) | 248 | 19 | 11 | 100 |
| Female | 3002 | 207 (6.9%) | 195 (6.5%) | 235 | 22 | 26 | 169 |
| **HLA-DR-DQ** |  |  |  |  |  |  |  |
| DR4-DQ8 only | 2483 | 69 (2.8%) | 41 (1.7%) | 228 | 3 | 3 | 38 |
| DR3-DQ2/DR4-DQ8 | 2366 | 157 (6.6%) | 92 (3.9%) | 170 | 18 | 9 | 83 |
| DR3/DQ2/DR3/DQ3 | 1303 | 172 (13.2%) | 173 (13.3%) | 85 | 20 | 25 | 148 |

<sup>a</sup>= Any child developing CDA and selected at random into sub-cohort serves as a time-varying control before CDA development.
<sup>b</sup>= CD-onset are children developing CDA before age 4-years and subsequently diagnosed with celiac disease.

**Table 4.** Case-cohort study designed from the TEDDY celiac cohort with CDA-hi ^b^ as an outcome of interest.

|  | TEDDY Celiac Cohort |  |  | MAVRiC Case-Cohort |  |  |  |
| --- | --- | --- | --- | --- | --- | --- | --- |
|  | Total<br>N | CDA-other<br>N<br>(% of total) | CDA-hi<br>N<br>(% of total) | Sub-cohort (random sample) |  |  | CDA hi<br>N <sup>b</sup> |
|  |  |  |  | Non CDA<br>N | CDA-other<br>N <sup>a</sup> | CDA hi<br>N <sup>ab</sup> |  |
| **Total** | 6132 | 502 (8.2%) | 202 (3.3%) | 483 | 53 | 25 | 177 |
| **Country** |  |  |  |  |  |  |  |
| US | 2472 | 159 (6.4%) | 71 (2.9%) | 190 | 17 | 9 | 62 |
| Finland | 1398 | 113 (8.1%) | 34 (2.4%) | 116 | 15 | 3 | 31 |
| Germany | 388 | 25 (6.8%) | 17 (4.6%) | 37 | 3 | 1 | 16 |
| Sweden | 1894 | 205 (10.8%) | 80 (4.2%) | 140 | 18 | 12 | 68 |
| **Sex** |  |  |  |  |  |  |  |
| Male | 3130 | 232 (7.4%) | 70 (2.2%) | 248 | 24 | 6 | 64 |
| Female | 3002 | 270 (9.0%) | 132 (4.4%) | 235 | 29 | 19 | 113 |
| **HLA-DR-DQ** |  |  |  |  |  |  |  |
| DR4-DQ8 only | 2483 | 79 (3.2%) | 31 (1.3%) | 228 | 6 | 0 | 31 |
| DR3-DQ2/DR4-DQ8 | 2366 | 183 (7.7%) | 66 (2.8%) | 170 | 19 | 8 | 58 |
| DR3/DQ2/DR3/DQ2 | 1303 | 240 (18.4%) | 105 (8.1%) | 85 | 28 | 17 | 88 |

<sup>a</sup>= Any child developing CDA and selected at random into sub-cohort serves as a time-varying control before CDA development
<sup>b</sup>= CDA-hi are children developing CDA before age 4 -years and tTGA level of the persistent sample was $\geq 60$ U/ml; Only 20 CDA-hi cases were not diagnosed with celiac disease during follow-up in TEDDY. Specificity for biopsy proven celiac disease within a year of seroconversion was >90%

Risk factors of CD-onset were re-examined on the case-cohort study. As expected, the month of birth between April and July as compared to other months (HR= 1.52, 95%CI = 1.11 – 2.08, p=0.008) and number of virus-related RIE reported August to October during the first (/additional report, HR= 1.23, 95%CI = 1.03 – 1.46, p=0.02) remained significantly associated with CD-onset after adjusting for the other risk factors. Similarly, April to June month of birth (HR= 1.93, 95%CI = 1.32 – 2.82, p=0.0006) as well as virus-related respiratory infections reported August to October (additional HR = 1.32, 95%CI = 1.08 – 1.63, p=0.008) were more strongly associated with CDA-hi.

## Discussion

Infections and gut microorganisms have been commonly implicated in the etiology or pathogenesis of celiac disease (13). However, the lack of large longitudinal studies among genetically at-risk populations has made it challenging to link exposures that vary with age and season, with a time-dependent specific risk of disease. TEDDY is the largest longitudinal study that has prospectively screened genetically at-risk populations in the US and Europe for the incidence of celiac disease autoimmunity before a celiac disease diagnosis. A retrospective examination of children with autoimmunity, with and without a subsequent biopsy-proven celiac disease diagnosis, has revealed virus-related respiratory infections during the months of August to October correlate specifically with a chronic form of autoimmunity leading to disease. Using these findings, we have presented a newly designed celiac disease case-cohort study that will provide an excellent opportunity to investigate the role of microbes prospectively by analyzing longitudinally collected blood and stool samples from enrolment at age 3-months until CDA develops. Analysis is currently ongoing to identify the celiac disease associated viruses by serology at regular 6-month intervals, by RT-PCR from 18214 monthly stool samples and through the analysis of the bacteriome to identify celiac disease changes associated in the whole microbiome.

Previous work in TEDDY has examined how reported infections correlate with the risk of a child developing celiac disease autoimmunity at an early age. However, only GIEs and not RIEs correlated with the risk of CDA (13). Despite studying 732 incident CDA cases in a total cohort population of 6,327, only GIEs reported in any 3-month period were significantly associated with an increased risk of CDA seroconversion by the next quarterly visit. Despite this, Kemppainen et al., did observe RIEs trending upwards 0-3 and 3-6 months prior to CDA seroconversion. Additionally, the magnitude of the correlation between GIE in a 3-month period and CDA seroconversion within the following 3-months was strongly dependent on other factors during the first year of life. This suggested the possibility of disease heterogeneity. In this study we excluded families whose child had developed diabetes autoimmunity before the age of 4 years. Additionally, we re-examined RIE and GIEs for correlations specifically for risk of celiac disease, i.e. CDA followed by biopsy-proven celiac disease diagnosis and, separately, children who developed CDA-only with no subsequent diagnosis. We found RIEs in any year, and particularly during the first year of life, specifically correlated with the onset of celiac disease. This difference suggests that RIEs as early as the first year of life conditions a more severe type of autoimmunity which may make it easier to identify respiratory type of viruses as a significant trigger of celiac disease among young children. Our findings adjusted for other strong TEDDY risk factors of CDA including HLA-DR3-DQ2, female predominance, non-HLA single nucleotide polymorphisms and a higher gluten intake during the first five years of life.^11^ A, small, nested metagenomics case-control study of fecal virome showed preliminary evidence of a cumulative effect of enterovirus infections and gluten on risk of CDA^12^. These results together suggest that the cumulative influence of infections during the fall season may be specific to a certain virus and may involve other factors that dictate how quickly CDA, and celiac disease develop.

Viruses that cause respiratory infections typically replicate on the respiratory mucosa. For example, the most common respiratory viruses, rhinoviruses, replicate mostly in the upper respiratory track, and some rhinovirus types also in lower respiratory track. However, some respiratory viruses can also replicate in the intestinal mucosa. The prime example is enteroviruses, which have associated with increased risk of celiac disease in previous studies. These viruses replicate both in the respiratory and intestinal mucosa, and can spread to the submucosal immune system, including gut-associated lymphoid tissue (GALT). Thus, they may interact with gluten in the intestinal mucosa and GALT, which offers one plausible mechanism how they could contribute to the initiation of CD-onset at an early age when gluten consumption is rapidly increasing. The most common symptom of enterovirus infection is common cold diseases while they only rarely cause gastroenteritis. Thus, in the present study which was based on questionnaire data enterovirus infections have mostly been classified as respiratory infection. They also peak at autumn time, in contrast to many other respiratory viruses. Another picornavirus, parechovirus, can also replicate in both respiratory and intestinal mucosa, and one prospective study has shown that detection of parechoviruses in stools was associated with increased risk of CD. However, the inflammatory immune response that leads to the destruction of small intestinal enterocytes and subsequent lesion may, in part, be driven by triggering agents such as enteroviruses along with bacteria. Enterovirus in the gut microbiome may present as pathogen-derived antigens and provoke inflammation early in life^6^. A similar mechanism has been observed in Epstein-Barr virus infection and its association with multiple sclerosis, where persistent infection acts as a driver of autoimmune reactions and, subsequently, disease development^7^.

The need to add additional CDA cases in the future and to stratify by other risk factors during analysis contributed to the decision to choose a case-cohort over a nested case-control study design. Nearly all children who develop celiac disease had started a GFD after CDA development. Moreover, none of the remaining CDA children with no diagnosis for celiac disease had started a GFD, **Supplemental Figure 4**. Most parents were not aware of their child’s CDA development until at least after the persistent sample. Less than 20% of children who did develop celiac disease autoimmunity and started a GFD were neither biopsied nor diagnosed with celiac disease. The reasons for starting a GFD diet were unknown.

Furthermore, it was possible for CD-onset children to be determined with low to medium tTGA levels (<60 U/ml) at CDA development and to be diagnosed with celiac diagnosis years later. It is unclear if these children had celiac disease at the time of CDA development. Our preliminary analysis suggests diet and genetic factors may play a stronger role. For this reason, we included a secondary outcome for sensitivity analysis and designed the MAVRiC study to better ensure any microbial associations found could be linked specifically and subsequently with onset of disease in young children and not because of reverse causation.

Results associated with CD-onset will be confirmed with CDA-high.

As mentioned above, children developing IA before 4 years of age were excluded. Most of these children developed IA during the first two years of life and parents learned of the results by the next quarterly visit. Of the 423 who developed IA, 50 (11.8%) developed CDA and 20/50 were confirmed with celiac disease. Similar viral and bacterial studies have been conducted in TEDDY and results may help investigate the impact of their exclusion from MAVRiC.

The first objective of the MAVRiC longitudinal study will be to identify enteroviruses and bacteria that correlate especially with the onset of celiac disease. Of importance is discovering these infections independent of known risk factors. Because of the large scale of TEDDY, its international scope across four countries on two continents, its standardized methods for screening CDA and for collection of samples, this project is expected to produce universal findings that will clarify the inconsistencies in the literature, enhance understanding of the celiac disease process, and provide compelling data for future mechanistic studies of viral or microbial triggers in autoimmunity. We hypothesize that the longitudinal impact of infections and microorganisms on the risk of disease will likely depend on the child’s age of exposure, time of disease onset and timing relative to age of gluten introduction. Further, identification of infectious triggers (or microbial dysbiosis) may lead to precise prevention strategies with vaccines and microbiome-enhancers.

In conclusion, evidence supports that the number of risk factors add up to increase risk of an earlier onset of celiac disease and the MAVRiC study will investigate the role of viruses as triggers at an early age with gut microbiome and diet as a time-dependent contribution.

## Data Availability

Data from The Environmental Determinants of Diabetes in the Young (https://doi.org/10.58020/y3jk-x087) reported here will be made available for request at the NIDDK Central Repository (NIDDK-CR) website, Resources for Research (R4R), https://repository.niddk.nih.gov/.

## Abbreviations

CDA: Celiac Disease Autoimmunity
GIE: gastrointestinal infectious episode
CI: confidence interval
HR: hazard ratio
RIE: respiratory infectious episode.

## Notes

**Conflict of Interest Disclosures**: DA receives consultant fees as member of Sanofís scientific advisory board. HH is a stock owner and member of the board of Vactech Oy which develops vaccines against picornaviruses.

**Grant Support**: This study was funded by the National Institute of Health (NIH/NIDDK 1 R01 DK124581-01). The TEDDY Study is funded by U01 DK63829, U01 DK63861, U01 DK63821, U01 DK63865, U01 DK63863, U01 DK63836, U01 DK63790, UC4 DK63829, UC4 DK63861, UC4 DK63821, UC4 DK63865, UC4 DK63863, UC4 DK63836, UC4 DK95300, UC4 DK100238, UC4 DK106955, UC4 DK112243, UC4 DK117483, U01 DK124166, U01 DK128847, and Contract No. HHSN267200700014C from the National Institute of Diabetes and Digestive and Kidney Diseases (NIDDK), National Institute of Allergy and Infectious Diseases (NIAID), Eunice Kennedy Shriver National Institute of Child Health and Human Development (NICHD), National Institute of Environmental Health Sciences (NIEHS), Centers for Disease Control and Prevention (CDC), and Breakthrough T1D (formerly JDRF). This work is supported in part by the NIH/NCATS Clinical and Translational Science Awards to the University of Florida (UL1 TR000064) and the University of Colorado (UL1 TR002535). The content is solely the responsibility of the authors and does not necessarily represent the official views of the National Institutes of Health.

### Competing Interest Statement

DA receives consultant fees as member of Sanofi's scientific advisory board. HH is a stock owner and member of the board of Vactech Oy which develops vaccines against picornaviruses. No other authors had a competing interest.

### Funding Statement

This study was funded by the National Institute of Health (NIH/NIDDK 1 R01 DK124581-01). The TEDDY Study is funded by U01 DK63829, U01 DK63861, U01 DK63821, U01 DK63865, U01 DK63863, U01 DK63836, U01 DK63790, UC4 DK63829, UC4 DK63861, UC4 DK63821, UC4 DK63865, UC4 DK63863, UC4 DK63836, UC4 DK95300, UC4 DK100238, UC4 DK106955, UC4 DK112243, UC4 DK117483, U01 DK124166, U01 DK128847, and Contract No. HHSN267200700014C from the National Institute of Diabetes and Digestive and Kidney Diseases (NIDDK), National Institute of Allergy and Infectious Diseases (NIAID), Eunice Kennedy Shriver National Institute of Child Health and Human Development (NICHD), National Institute of Environmental Health Sciences (NIEHS), Centers for Disease Control and Prevention (CDC), and Breakthrough T1D (formerly JDRF). This work is supported in part by the NIH/NCATS Clinical and Translational Science Awards to the University of Florida (UL1 TR000064) and the University of Colorado (UL1 TR002535). The content is solely the responsibility of the authors and does not necessarily represent the official views of the National Institutes of Health.

### Author Declarations

Colorado Multiple Institutional Review Board, Georgia Medical College of Georgia Human Assurance Committee (2004-2010)/Georgia Health Sciences University Human Assurance Committee (2011-2012)/Georgia Regents University Institutional Review Board (2013-2016)/Augusta University Institutional Review Board (2017-present), University of Florida Health Center Institutional Review Board, Washington State Institutional Review Board (2004-2012)/Western Institutional Review Board (2013-2019)/WCG IRB (2020-present), and European Ethics Committee Boards of Finland Ethics Committee of the Hospital District of Southwest Finland, Germany Bayerischen Landesärztekammer (Bavarian Medical Association) Ethics Committee, Sweden Regional Ethics Board in Lund, Section 2 (2004-2012)/Lund University Committee for Continuing Ethical Review (2013-present) gave ethical approval for this work.

