## Supplement Table and Figures for "A case-cohort longitudinal study for the analysis of microbial associations and viruses on the risk of celiac disease (MAVRiC)"

**Supplemental Figure 1**

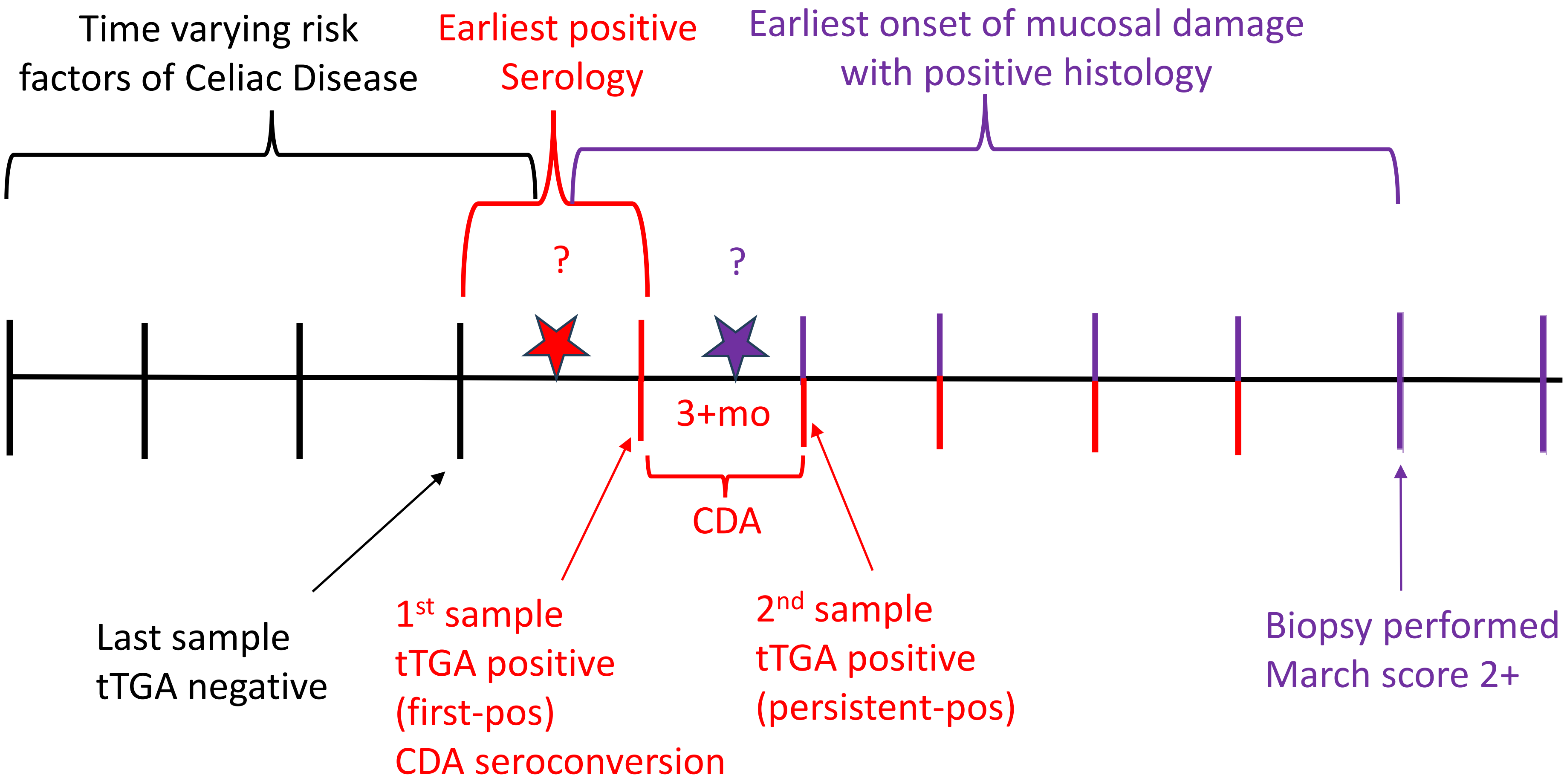


CD-onset = CDA seroconversion age < 4years followed by a positive biopsy

CD-hi = CDA seroconversion age <4 years with titer of persistent-positive sample ≥60 U/ml

#### Supplemental Figure 2

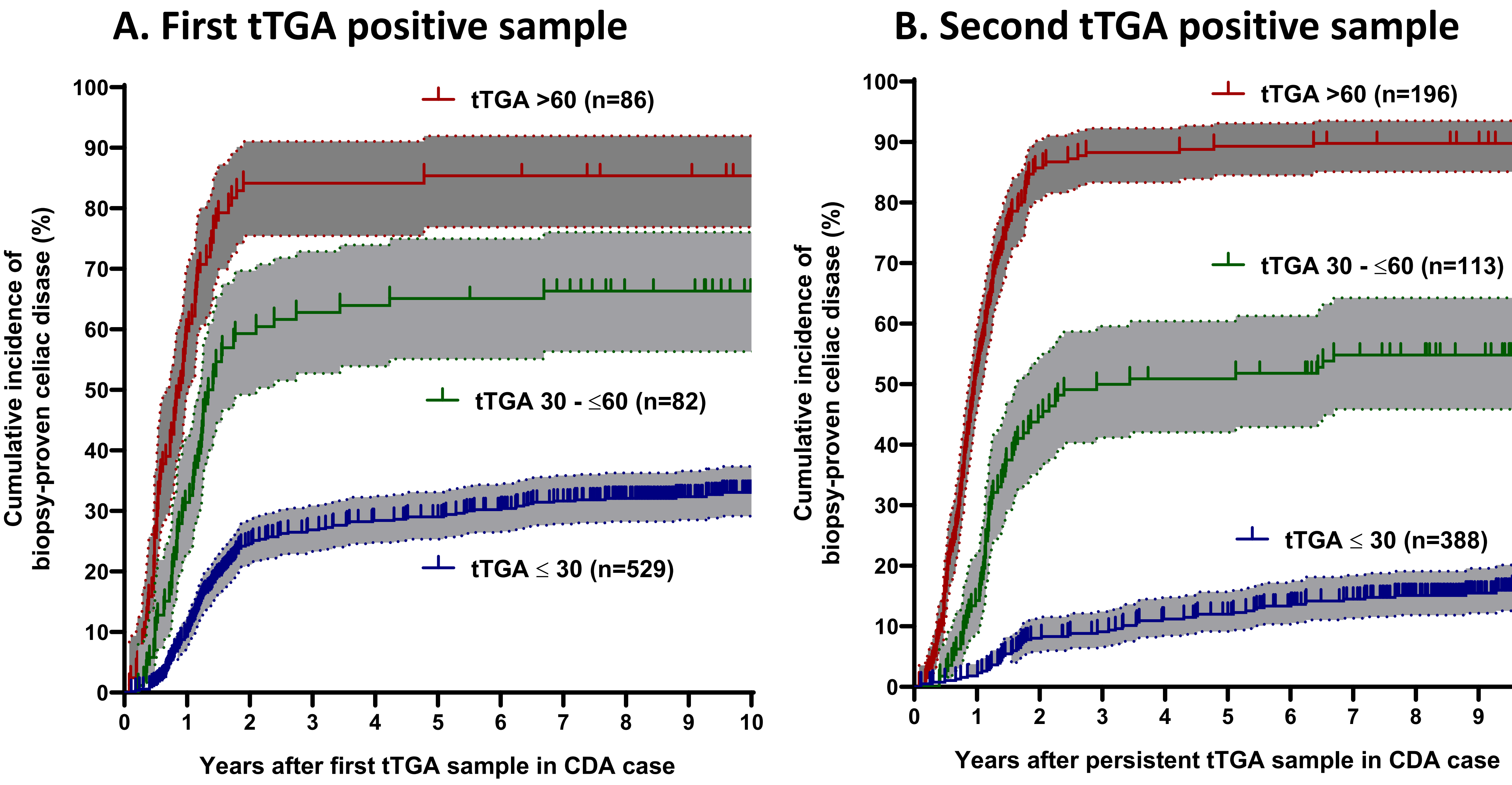

Supplemental Figure 3

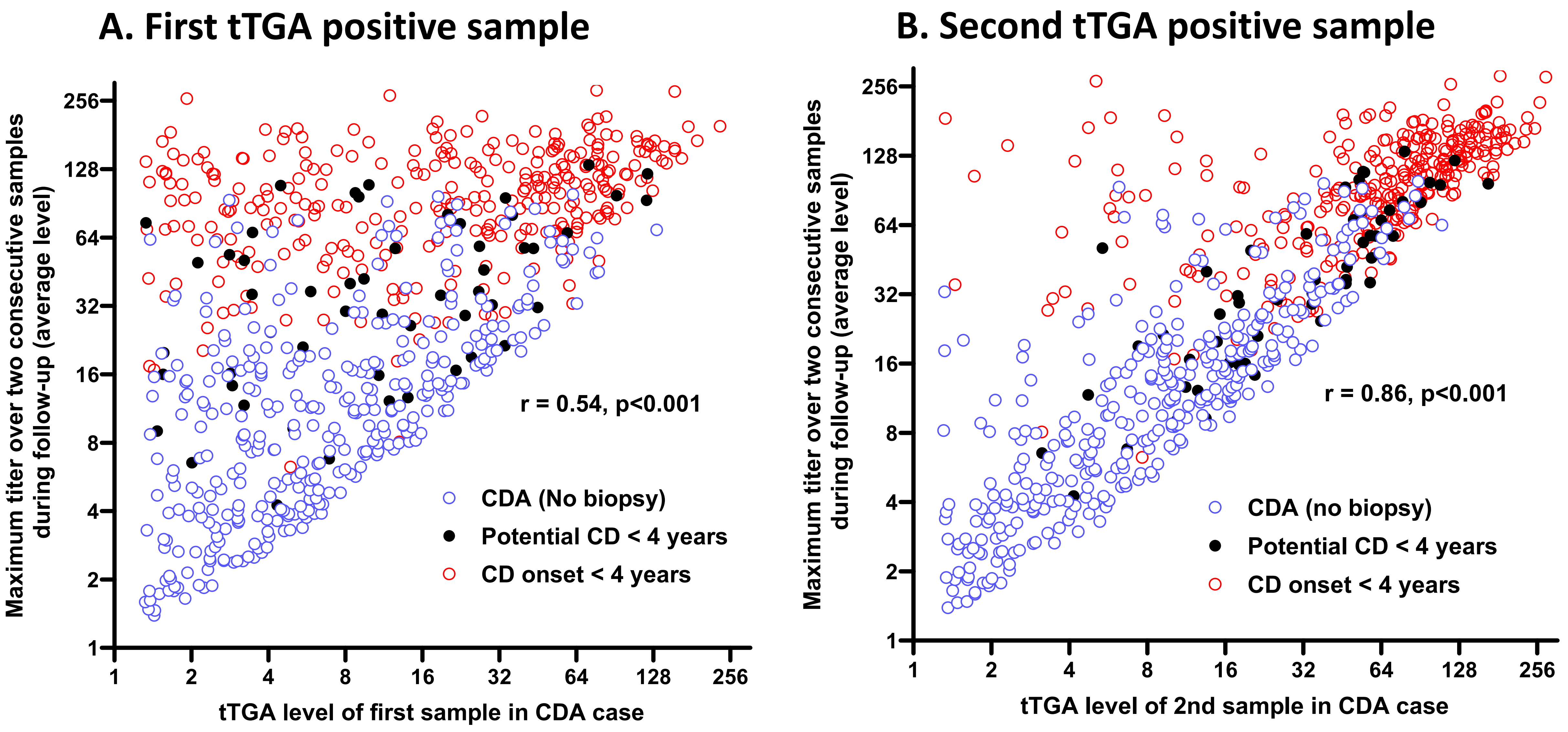

### Supplemental Figure 4

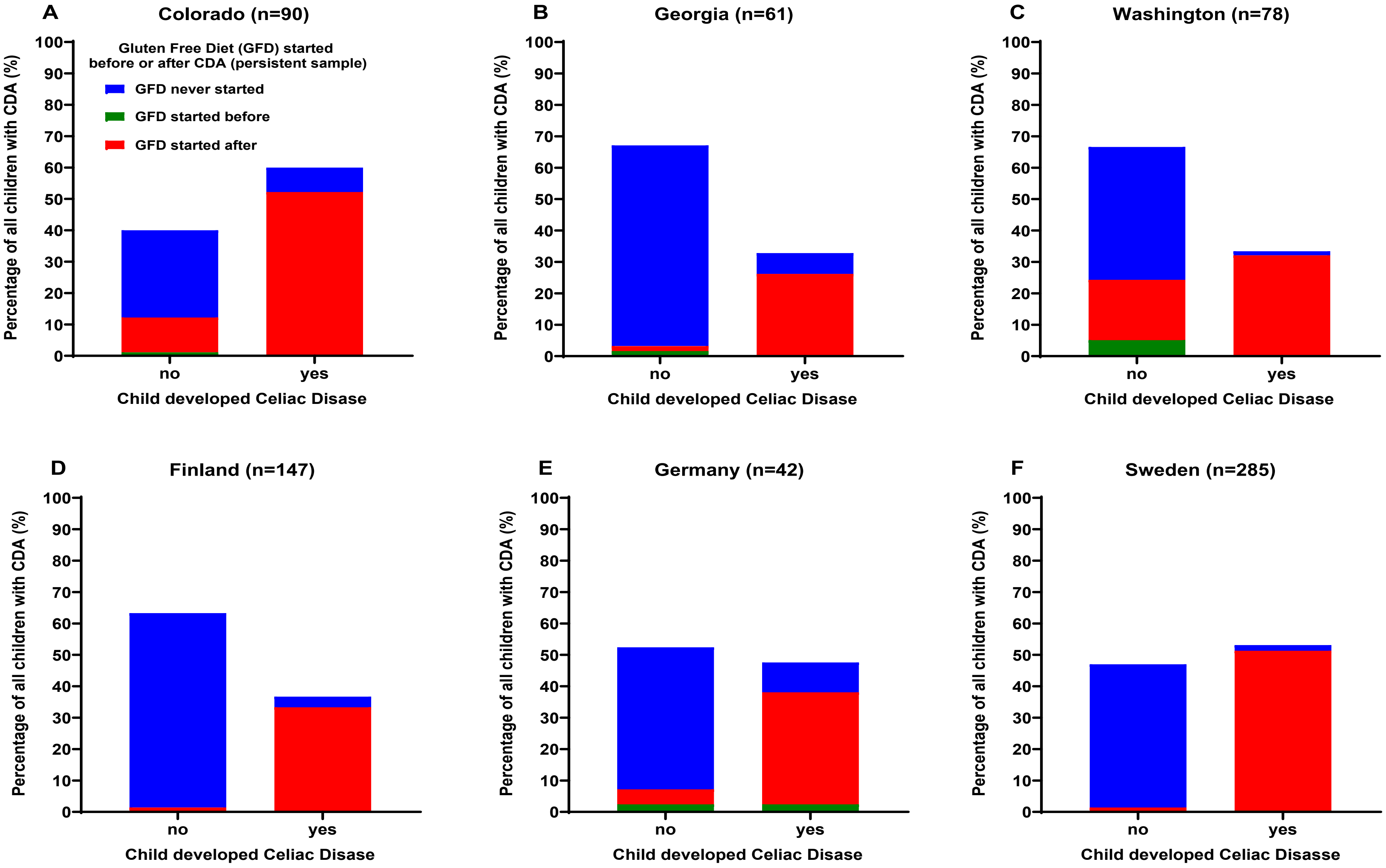

**Supplemental table 1** SNPs used in the calculation of a non-HLA polygenic risk score (PRS) for the onset of celiac disease (CD-onset) before age 4-years.

| SNP | Chromosome | Position (GRCh38) | Minor Allele [a] | Nearby Gene | log HR [b] (CD-onset) |
|---|---|---|---|---|---|
| rs72704176 | 1 | 155456067 | T | ASHIL | 0.5831 |
| rs115195008 | 1 | 197649315 | C | DENND1B | 0.5324 |
| rs3771689 | 2 | 159365291 | T | BAZ2B | -0.3202 |
| rs1829618 | 2 | 211568600 | A | ERRB4 | 0.2669 |
| rs12990970 | 2 | 203835966 | T | CTLA-4 | -0.1816 |
| rs13014907 | 2 | 185208879 | T | ZNF804A | 0.7024 |
| rs1464510 | 3 | 188394766 | A | LPP | 0.2671 |
| rs6806528 | 3 | 69203748 | T | FRMD4B | 0.3886 |
| rs12493471 | 3 | 45910186 | T | CCR9 | 0.2200 |
| rs114157400 | 4 | 102014304 | G | BANK1 | 0.4529 |
| rs1054091 | 6 | 159048480 | C | TAGAP | 0.3737 |
| rs2327832 | 6 | 137651931 | G | TNFAIP3 | 0.3137 |
| rs802734 | 6 | 127957653 | G | PTPRK | 0.2901 |
| rs61751041 | 7 | 107953740 | T | LAMB1 | 0.6360 |
| rs6967298 | 7 | 70549523 | G | AUTS2 | -0.3899 |
| rs2409747 | 8 | 11220953 | T | XKR6 | 0.3830 |
| rs76554494 | 9 | 120628467 | C | MEGF9 | 0.3314 |
| rs9423406 | 10 | 5311727 | A | AKR1C7P | 0.2752 |
| rs117561283 | 12 | 68052813 | T | IFNG | 0.5034 |
| rs653178 | 12 | 111569952 | C | SH2B3 | 0.1791 |
| rs8013918 | 14 | 75242863 | T | FOS | -0.2405 |
| rs11203203 | 21 | 42416077 | A | UBASH3A | 0.2157 |
| rs2298428 | 22 | 21628603 | T | YDJC | 0.3624 |

[a] = Forward strand based on the Genome Reference Consortium Human Build 38.

[b] = The PRS is created by summing across single nucleotide polymorphisms (SNPs) the product of the number of minor alleles by the log beta coefficient (log hazard ratio=log HR) that was estimated from a multivariate proportional hazard model on specific risk of CD-onset adjusting for country, number of HLA-DR3-DQ2 haplotypes, sex and first three principal components describing ancestry.
